# Beyond Single Biomarkers: A Graph Neural Network Framework for Multivariable Prediction of Clinical Outcomes from Brain Imaging

**DOI:** 10.64898/2026.06.21.26356202

**Authors:** Jamal Esmaelpoor, Abdolrahim Kadkhodamohammadi, Tommy Peng, Beth Jelfs, Darren Mao, Abdollah Ghafouri, Maureen J. Shader

**Affiliations:** Bionics Institute, Fitzroy, VIC 3065, Australia; Department of Medical Bionics, University of Melbourne, Parkville, VIC 3010, Australia; School of Engineering, Swinburne University of Technology, Hawthorn, VIC 3122, Australia; Surgical Research and Technology, Medtronic, London EC1V 2QY, U.K; Department of Electronic, Electrical and Systems Engineering, University of Birmingham, Birmingham B15 2FG, U.K.; Islamic Azad University, Boukan Branch, Boukan 159, Iran; Department of Speech, Language, and Hearing Sciences, Purdue University, West Lafayette, IN 47907, USA

**Keywords:** Brain imaging, brain networks, regional brain activation, functional near-infrared spectroscopy, graph neural networks, multivariable analysis, outcome prediction

## Abstract

Understanding brain–behavior relationships requires models capturing the distributed, interactive, and multiscale nature of neural systems. Traditional univariate approaches and single-biomarker models are inherently limited in this context, as they fail to represent dependencies across regions and the hierarchical organization of brain networks. In this study, we propose a graph-based multivariable framework for brain imaging analysis that integrates key organizational principles of brain function—including segregation, integration, modularity, and temporal dynamics—within a unified graph neural network architecture. The framework represents brain data as hierarchical graphs, where node features encode regional activation and temporal variability, and graph structure captures interactions within and between functional modules. The proposed approach is evaluated using functional near-infrared spectroscopy (fNIRS) data as a case study, where subject-specific brain graphs are constructed from task-based recordings acquired shortly after cochlear implant activation to predict speech understanding outcomes one year later. Under leave-one-subject-out validation, the model demonstrates strong predictive performance (R = 0.73, p *<* 0.001), outperforming previously reported single-biomarker approaches. Perturbation-based analyses further show that predictions are driven by distributed patterns of activity and interaction across regions and modalities, rather than isolated features. These results illustrate the capability of the proposed framework to capture complex brain organization and highlight its potential as a generalizable platform for multivariable analysis and prediction in neuroimaging applications beyond the specific clinical use case considered here.

## I. Introduction

THE human brain is a highly complex and adaptive system in which function arises from the coordinated activity of distributed regions and their interactions over time. Cognitive processes such as perception, language, and working memory are not localized to isolated anatomical areas; rather, they emerge from dynamic patterns of communication between specialized regions [1]. Accordingly, brain function is inherently distributed and interactive, and studying individual regions or single metrics provides only a partial and incomplete description of neural processes. This organization is further characterized by a modular structure, where groups of regions form functionally specialized subsystems. However, effective behavior depends not only on processing within these modules but also on interactions between them, requiring a balance between within-module specialization and between-module integration [2].

These principles are reflected in brain connectivity, which provides a high-dimensional representation of interactions across regions. The number of possible connections increases rapidly with the number of regions, and these features are strongly interdependent, exhibiting coordinated variation across the network. As a result, changes in brain function are rarely confined to individual regions or connections but instead manifest as distributed patterns spanning multiple components of the system [3]. Such network-level organization highlights that meaningful brain signals are embedded in structured interactions rather than isolated features. These characteristics impose fundamental limitations on traditional univariate approaches. By treating features independently, such methods ignore dependencies between variables, fail to capture distributed effects, and suffer from multiple comparison constraints that reduce statistical power. Consequently, meaningful system-level changes may remain undetected when analysis is restricted to isolated biomarkers or connections [4], [5]. In this context, focusing on individual features in isolation is analogous to searching for a signal in noise, as the underlying effects are distributed across coordinated patterns rather than localized to single elements.

Multivariable approaches provide a principled framework to address these challenges by modeling multiple features jointly and capturing their dependencies. By considering distributed patterns of information rather than isolated features, these methods offer the potential to better accommodate the high dimensionality and interdependence inherent in brain imaging data. When combined with predictive modeling and cross-validation, they also provide a more meaningful evaluation of whether brain-derived features generalize to unseen individuals [4], [5]. Taken together, these considerations highlight the need for hypothesis-free, multivariable frameworks that explicitly model interactions, dependencies, and modular organization in brain data, thereby enabling a more faithful representation of brain function and its relationship to behavior.

Recent advances in neuroimaging analysis have increasingly adopted multivariable, data-driven approaches to capture the distributed nature of brain function. Early studies demonstrated that high-dimensional functional connectivity patterns can be used for subject-level prediction using multivariate learning frameworks such as support vector machines [6]. Subsequent work extended these ideas by incorporating task-dependent and dynamic connectivity, showing that patterns of interaction across regions encode cognitive states and behavioral variability [7]. More recently, deep learning approaches have been applied to brain imaging data, including convolutional neural networks operating on volumetric images [8], [9] and graph-based models designed to capture relational structure and temporal dynamics [10]. Despite these advances, several limitations remain. Many multivariable approaches rely on fixed-size representations derived from predefined connectivity measures, such as correlation, which impose assumptions about the nature of interactions and often ignore their nonlinear and context-dependent characteristics [4], [6]. Image-based deep learning models, while powerful, treat the brain as a regular grid and do not explicitly model its network organization. Graph-based approaches address this limitation but frequently depend on large, noisy networks and complex architectures, which can reduce interpretability and limit their applicability in clinical settings. In addition, node representations in these models are often derived directly from connectivity profiles, leading to redundancy between node features and graph structure and limiting physiological interpretability.

In this study, we propose a graph-based multivariable framework to model key organizational principles of brain function, including segregation, integration, modularity, and temporal dynamics. The approach is implemented using a graph neural network (GNN), where node features capture regional brain activation in response to task conditions, reflecting functional segregation, alongside features describing trial-to-trial variability to account for dynamic aspects of neural responses. The input graphs are constructed to explicitly incorporate the hierarchical modular structure of the brain, enabling representation of both within-module organization and inter-module interactions. Through message passing, the GNN integrates information from neighboring nodes to learn higher-level representations that reflect both local activity and network context. The proposed framework is evaluated using functional near-infrared spectroscopy (fNIRS) recordings acquired shortly after cochlear implant (CI) activation, with the aim of predicting speech understanding outcomes one year post-implantation. Model performance is assessed using subject-level validation and compared against previously reported univariate approaches applied to similar datasets. In addition, we perform exploratory analyses to examine model behavior and provide insights into the relative contribution of different components of the multivariable representation.

## II. Methods

We propose a graph-based multivariable framework to model brain imaging data by integrating multiple neurobiological principles, including regional activation, inter-regional connectivity, and temporal dynamics. In this work, the framework is instantiated using fNIRS recordings from CI users, where each subject is represented as a graph in which node features are derived from task-based responses, and edges are defined based on the underlying modular organization of the brain network.

### A. Participants

We enrolled 31 postlingually deafened adult CI recipients shortly after implantation (mean age: 58.9 *±* 13.8 years; 19 males, 12 females). Ethical approval for the study was obtained from the Royal Victorian Eye and Ear Hospital (approval no. 19.1418H), and all participants provided written informed consent.

Given the relatively older age of the cohort, cognitive function was assessed to exclude potential cognitive decline. Cognitive performance was evaluated using the Trail Making Test Parts A and B [11]. In Part A, participants were required to connect numbered circles in ascending order, while in Part B they alternated between numbers and letters in sequence. Both tasks were administered on paper with randomly distributed targets, and participants were instructed to complete them as quickly and accurately as possible without lifting the pencil. All participants demonstrated performance within the normal range according to established criteria [12].

CI outcomes were assessed one year after device activation using a speech perception task. Participants completed a Bamford–Kowal–Bench (BKB) sentence test consisting of 15 sentences presented in four-talker babble noise at a signal-to-noise ratio of +10 dB [13]. Speech stimuli were delivered in free field at 65 dBA. To isolate performance with the implant, the contralateral ear was occluded and masked when appropriate. Performance was quantified as the percentage of correctly repeated words.

### B. Experimental Paradigm

The study employed a task-based design incorporating both audio-only and visual-only speech conditions, as described by [14]. Participants were seated comfortably in a sound-attenuated booth, facing a monitor, with a loudspeaker positioned one meter away for auditory stimulus delivery at 65 dBA under free-field conditions. The experimental protocol consisted of 36 contextually continuous segments from the children’s story Mrs. Tittlemouse by Beatrix Potter, narrated by a female native speaker of Australian English. These segments were presented across 18 audio-only and 18 visual-only trials (Figure 1c). In addition, 10 control trials were included, during which participants fixated on a gray cross in the absence of auditory or visual speech stimuli. Each segment had an average duration of 12.5 s, followed by an inter-trial interval of 15–30 s, during which participants maintained fixation on a gray cross in silence. To minimize order effects, trials were pseudorandomized across conditions while preserving the narrative continuity of the story within each modality. Brain imaging data acquired during the audio and visual conditions were used to construct the GNN model.

**Fig. 1.**
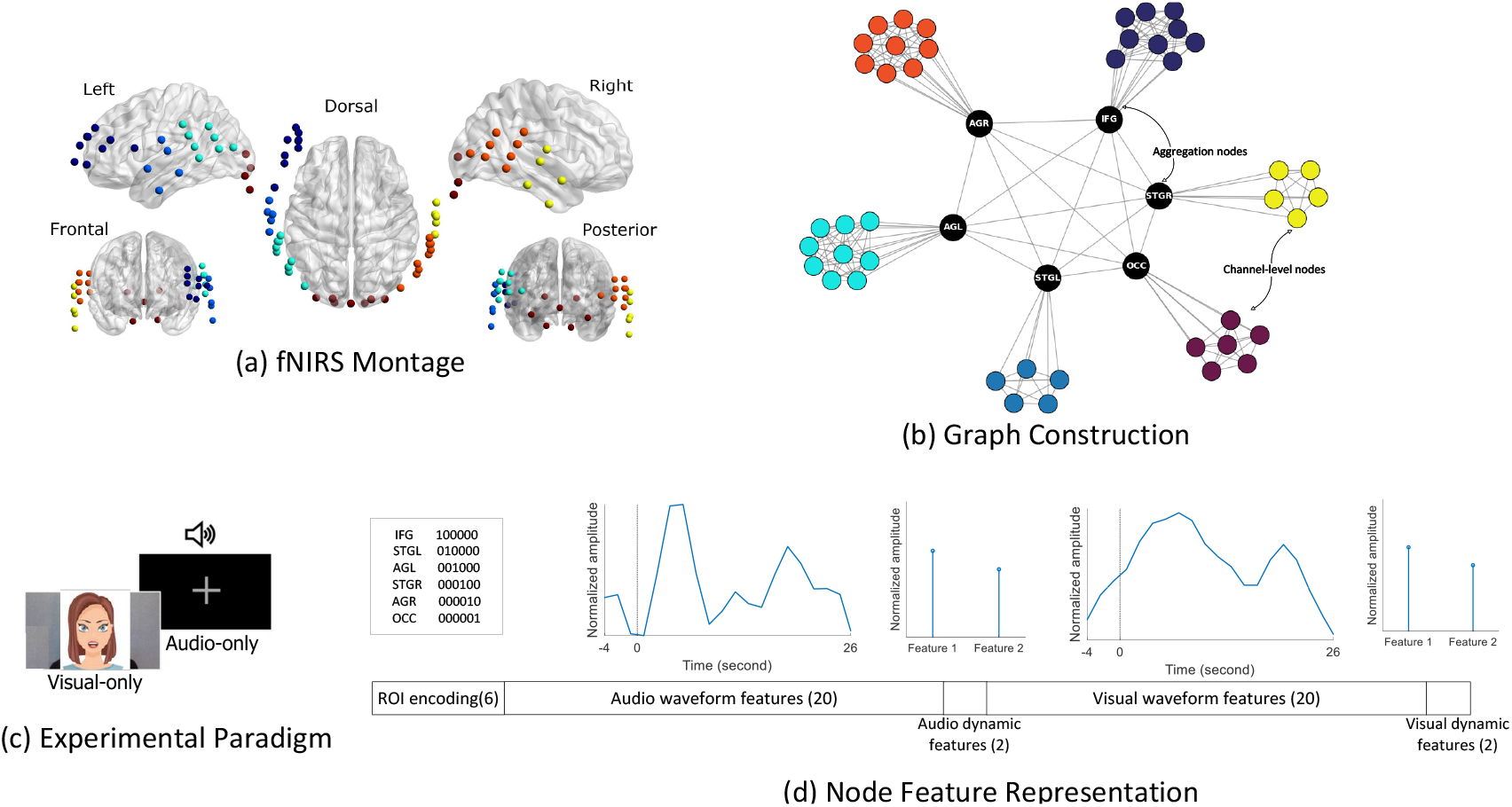
Multivariable graph-based representation of fNIRS data. (a) fNIRS optode montage showing spatial distribution of measurement channels across cortical regions, including frontal, temporal, and occipital areas. (b) Graph construction illustrating the hierarchical representation of brain networks. Channel-level nodes (colored) within each ROI are densely connected to model intra-regional interactions, while ROI-level aggregation nodes (black) summarize regional activity and are interconnected to capture inter-regional relationships. (c) Experimental paradigm consisting of audio-only and visual-only speech conditions. Participants were presented with segments of continuous speech in either modality, enabling the extraction of task-specific brain responses. (d) Node feature representation. Each node is described by a multivariable feature vector including ROI encoding, average hemodynamic response waveforms for audio and visual conditions, and dynamic features capturing trial-to-trial variability. Aggregation nodes contain ROI-level summaries of these features.

### C. fNIRS Data Acquisition

fNIRS recordings were conducted within the first month following CI activation to assess cortical activity. Data were acquired using a continuous-wave NIRScout system (NIRScout, NIRx Medical Technologies, LLC). The optode montage comprised 16 sources and 16 photodetectors, with each source emitting near-infrared light at dual wavelengths of 760 nm and 850 nm. This configuration yielded 44 long channels with an average source–detector separation of approximately 3 cm. The montage covered key cortical regions for visual and auditory language processing, including the inferior frontal gyrus, bilateral auditory cortices, and the occipital lobe (Figure 1a). To reduce contamination from extracerebral physiological signals (e.g., scalp blood flow), eight short-separation channels (8*mm*) were included. These channels were used to capture superficial hemodynamic activity, facilitating improved isolation of cortical signals [15]. For clarity, short-separation channels are not shown in Figure 1a.

### D. Data Preprocessing

Data preprocessing was performed using MATLAB and the NIRS Toolbox [16]. Raw fNIRS signals were first converted to optical density, and channel quality was assessed using the scalp coupling index (SCI) [17]. Channels with SCI values below 0.4 were considered unreliable and excluded from further analysis. To mitigate motion artifacts, the temporal derivative distribution repair method was applied [18]. The optical density signals were then converted to oxyhemoglobin (HbO) and deoxyhemoglobin (HbR) concentration changes using the modified Beer–Lambert law [19].

Because long-separation channels capture both cortical signals and systemic physiological noise (e.g., cardiac pulsation, respiration, and Mayer waves), additional steps were taken to isolate neural activity. Short-channel regression was performed by removing signals recorded from short-separation channels, which predominantly reflect superficial physiological fluctuations, from the long-channel measurements [20]. A Butterworth band-pass filter (8th order, 0.015–0.20 Hz) was subsequently applied to attenuate residual low-frequency drift and high-frequency physiological noise.

The cleaned hemodynamic signals were then epoched from − 4 to 26.3 s relative to stimulus onset and baseline-corrected using the pre-stimulus interval (− 4 to 0 s). Following preprocessing, the epoched data (original sampling rate: 3.91 Hz) were downsampled by a factor of 6. This step was performed to reduce temporal redundancy and computational load, as the frequency content of interest was well below the Nyquist limit after band-pass filtering [21]. All subsequent analyses were conducted using HbO signals, as prior studies have shown that HbO provides more robust and reliable measures of functional connectivity compared to HbR [22].

### E. Graph Representation of Brain Networks

#### 1) graph construction

To construct the graph representation, we first defined modules by selecting anatomically and functionally relevant regions of interest (ROIs) (Figure1a). The left frontal region was included to capture activity in the left inferior frontal gyrus, a key area associated with speech and language processing [23]. Following the approach of [14], the bilateral auditory cortices were further subdivided into two subregions: the superior temporal gyrus (STG) and the angular gyrus (AG). These regions—referred to as Heschl’s gyrus and planum temporale in their study—represent distinct anatomical components of primary and secondary auditory processing, respectively. Finally, a visual region encompassing the occipital lobe was defined to capture cortical responses to speech stimuli [24].

To capture the hierarchical organization of brain networks, we constructed the graph in a two-level manner reflecting both intra-regional and inter-regional interactions (Figure 1b). At the first level, nodes corresponding to fNIRS channels within each ROI were densely connected, enabling the model to capture local interactions and functional relationships within individual cortical regions. At the second level, we introduced a representative node for each ROI, referred to as an *aggregation node*, which summarizes information from all channels within that region. These aggregation nodes were then connected to each other, forming a higher-level graph that models interactions between different brain regions. This hierarchical design allows the graph to simultaneously represent fine-grained local connectivity within regions and coarse-grained interactions across regions.

#### 2) Node Feature Representation

Each node in the graph was assigned a feature vector designed to capture complementary aspects of brain activity, including spatial identity, task-evoked responses, and temporal dynamics (Figure 1d). For channel-level nodes, the feature vector comprised three components: (i) a one-hot encoding indicating the ROI membership of the channel, (ii) waveform features derived from the average hemodynamic responses during the audio-only and visual-only conditions (20 time points per condition), with the resulting waveforms globally z-normalized to ensure comparability across subjects and channels, and (iii) dynamic features (two per condition) characterizing trial-to-trial temporal variations.

The dynamic features were designed to capture both changes in response amplitude over time and the consistency of responses across trials. The first feature quantified the difference in peak amplitude between early and late trials, computed as the difference between the maximum of the average waveform across the first four trials and that across the last four trials. The second feature captured temporal consistency by computing the mean correlation between successive averaged trial groups. Specifically, trials were grouped into consecutive sets of three, the average waveform was computed for each group, and Pearson correlations were calculated between successive group averages; the mean of these correlations was then used as the feature.

For aggregation nodes, which represent ROI-level summaries, features were constructed by aggregating information across all channels within the corresponding region. The waveform features were computed as the average response across channels within each ROI for both audio and visual conditions. The dynamic features were defined as the average of the corresponding dynamic features extracted from all channel-level nodes within the ROI. The ROI encoding for aggregation nodes was set to a zero vector (000000), allowing the model to distinguish them from channel-level nodes while preserving a consistent feature dimensionality.

#### 3) Data Augmentation Strategy

To improve model robustness and mitigate the limited sample size, a two-stage data augmentation strategy was employed to generate multiple graph instances per subject. First, trial-level variability was introduced by randomly removing one trial from the set of available trials for each condition. This procedure was repeated nine times, in addition to the original full-trial dataset, resulting in 10 augmented versions capturing variations in trial composition. Second, channel-level variability was incorporated by randomly removing one or two channels from each of the previously generated graphs. This step was repeated four times per graph, along with the original version, resulting in five variants per trial-augmented graph.

### F. Graph Attention Network for Outcome Prediction

To predict clinical outcomes from the constructed brain graphs, we employed a graph attention network (GAT) architecture that integrates node-level features with graph topology (Fig. 2) [25]. The model was implemented using the PyTorch Geometric framework [26]. Each subject-specific graph was processed independently, where nodes represent fNIRS channels and ROI-level aggregation units, and edges encode functional relationships between them.

**Fig. 2.**
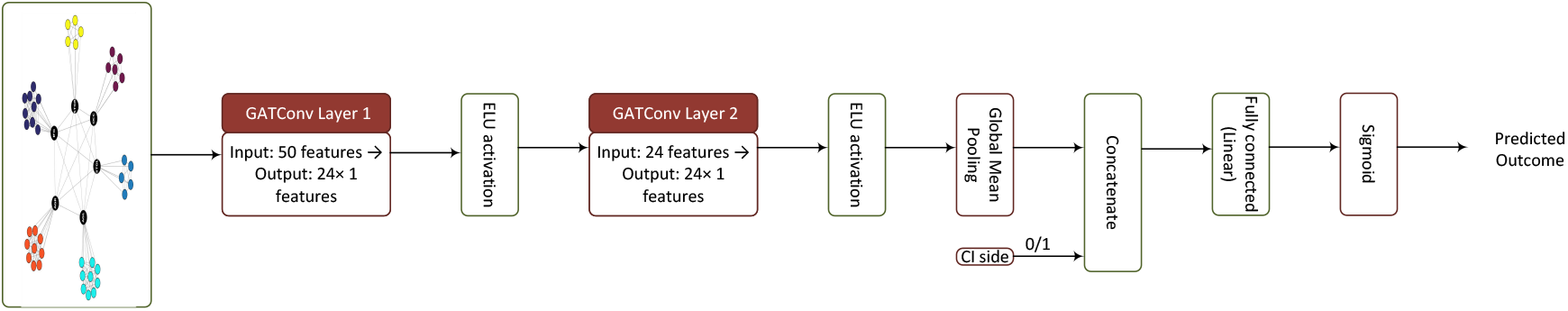
Graph attention network architecture for outcome prediction. The model processes subject-specific brain graphs through two successive graph attention convolution layers (GATConv), each followed by an ELU activation. These layers learn feature-dependent, weighted interactions between neighboring nodes via attention-based message passing. Node embeddings are then aggregated using global mean pooling to produce a graph-level representation. A binary variable indicating cochlear implant side is concatenated with this representation prior to a fully connected layer and sigmoid activation, yielding a normalized prediction of speech understanding outcome.

The network consists of two successive graph attention convolution layers (GATConv), each followed by an exponential linear unit (ELU) activation function. The first layer maps the input node features to a 24-dimensional embedding space, enabling the model to learn weighted combinations of neighboring node features through attention-based message passing. The second GAT layer further refines these representations, producing updated node embeddings that capture higher-order interactions within the graph.

Following the convolutional layers, a global mean pooling operation is applied to aggregate node embeddings into a single graph-level representation for each subject. This step enables the model to summarize information across both channel-level and aggregation-level nodes. In addition to the graph-derived features, a binary variable indicating CI side (left/right) is incorporated as an auxiliary input. This variable is concatenated with the pooled graph representation prior to the final prediction stage. The resulting feature vector is passed through a fully connected (linear) layer, followed by a sigmoid activation function to produce the final prediction. The model outputs a normalized (0–1) estimate of speech understanding performance, where the original speech scores range from 0 to 100%.

Model training was performed using the Adam optimizer with a learning rate of 2.5 *×* 10^−4^ and weight decay of 5 *×* 10^−4^. A Huber loss function was used to provide robustness to potential outliers in the target values [27]. The learning rate was reduced during training using a step decay schedule. To prevent overfitting, early stopping was implemented based on the training loss with a predefined patience criterion.

Model performance was evaluated using a leave-one-subject-out (LOSO) cross-validation scheme. In each fold, all graphs corresponding to one subject were held out for testing, while the remaining subjects were used for training. Predictions for each subject were obtained by averaging the outputs across all augmented graph instances associated with that subject.

## III. Results

### A. Performance of the Proposed GNN Model in Predicting Speech Outcomes

As shown in Figure 3a, the proposed model demonstrated a strong correspondence between predicted and true scores, with a Pearson correlation of *R* = 0.73(*p <* 0.001). The regression line closely followed the identity line, indicating that the model was able to capture inter-subject variability in speech performance with good fidelity. Importantly, the model maintained consistent performance across the full range of outcome scores, suggesting robustness to variability in both lower- and higher-performing individuals. The use of LOSO validation provides a stringent evaluation of generalization performance, as the model is trained exclusively on data from other subjects and evaluated on a completely unseen subject.

**Fig. 3.**
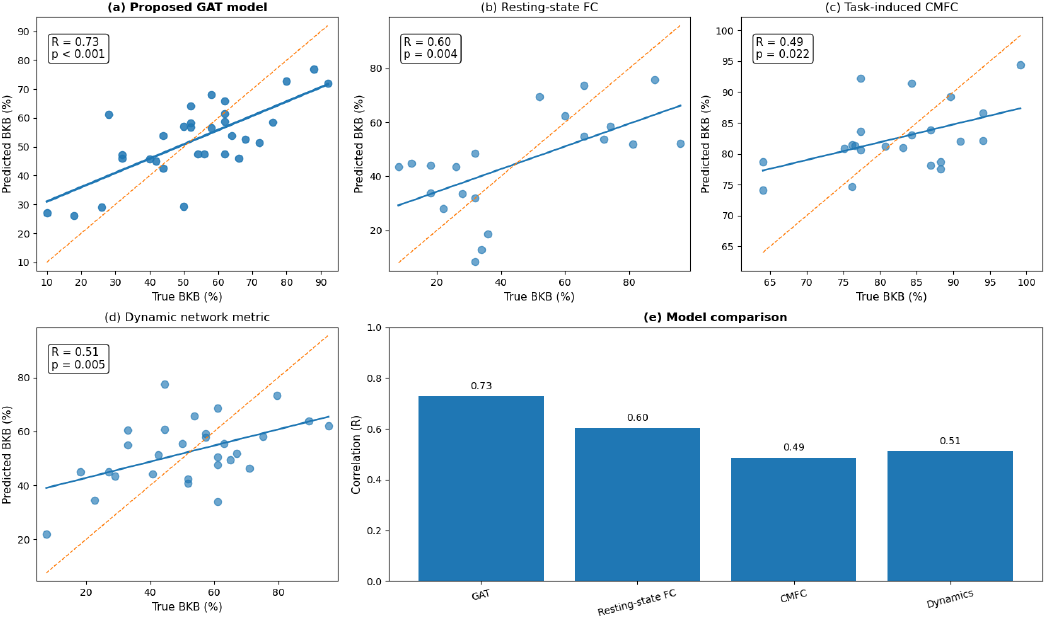
Predictive performance of the proposed GNN model and comparison with single-biomarker approaches. (a) Scatter plot of predicted versus true speech understanding scores for the proposed GNN model under LOSO validation. The solid line indicates the fitted regression, and the dashed line represents the identity line. (b–d) Corresponding regression plots for three single-biomarker approaches: resting-state functional connectivity, task-induced cross-modal functional connectivity, and dynamic functional connectivity. (e) Summary comparison of predictive performance across models, shown as Pearson correlation coefficients between predicted and true outcomes.

### B. Comparison with Single-Biomarker Approaches

To contextualize the performance of the proposed multivariable framework, we compared its predictive capability with three previously reported single-biomarker approaches derived from our earlier studies (Figure 3b-d). These approaches are hypothesis-driven and each relies on a single neurophysiological indicator associated with CI outcomes. The first approach is based on resting-state functional connectivity, where the average clustering coefficient serves as a proxy for network organization [28]. This metric has been interpreted as reflecting neural degradation following hearing loss and subsequent restoration after implantation, and has been linked to speech understanding outcomes. The second approach focuses on task-based cross-modal functional connectivity (CMFC), quantifying contralateral audio–visual functional connectivity relative to the implanted side as an indicator of speech performance [29]. The third approach examines dynamic functional connectivity, using the network switching rate as a measure of temporal instability in brain modular organization, which has also been shown to correlate with CI outcomes [30]. To ensure a fair comparison, all single-biomarker models were re-evaluated using a LOSO validation framework, consistent with the evaluation strategy adopted in the current study. For each biomarker, a regression model was trained on all subjects except one, and the speech score of the held-out subject was estimated solely based on the corresponding biomarker value. This procedure provides a more stringent and realistic assessment of predictive performance compared to the simple correlation analyses typically reported in prior work. The results shown in Figure 3 indicate that, although each biomarker maintains a meaningful association with speech outcomes, their predictive performance under subject-level validation is limited compared to the proposed GNN model.

### C. Exploratory Analyses of Model Behaviour

To investigate the behavior of the proposed model, we performed a series of perturbation-based analyses to quantify the contribution of different components of the input graph. In this framework, specific elements of the node features or graph structure were systematically modified, and the resulting change in the model’s prediction was measured [31].

For feature-level analysis, the contribution of individual feature groups was assessed by replacing the corresponding feature values with neutral inputs while keeping the rest of the graph unchanged. Continuous features (e.g., waveform and dynamic features) were replaced with zero-valued inputs following normalization, whereas binary features (e.g., ROI encoding) were masked accordingly. The importance of each component was then quantified as the absolute change in the predicted outcome relative to the original prediction.

These perturbations were applied across different levels of the graph representation, including feature groups, node attributes, and connectivity patterns. The resulting importance measures reflect the sensitivity of the model to specific inputs and are interpreted in a relative sense to compare the contribution of different components. Among the range of analyses performed, we focus here on those that provided the most informative and interpretable insights into model behavior.

### D. Node-Level Feature Importance Analysis

To investigate the relative contribution of different components of the node feature vector, we performed a perturbation-based feature importance analysis at both the feature-block and individual-feature levels (Figure 4).

**Fig. 4.**
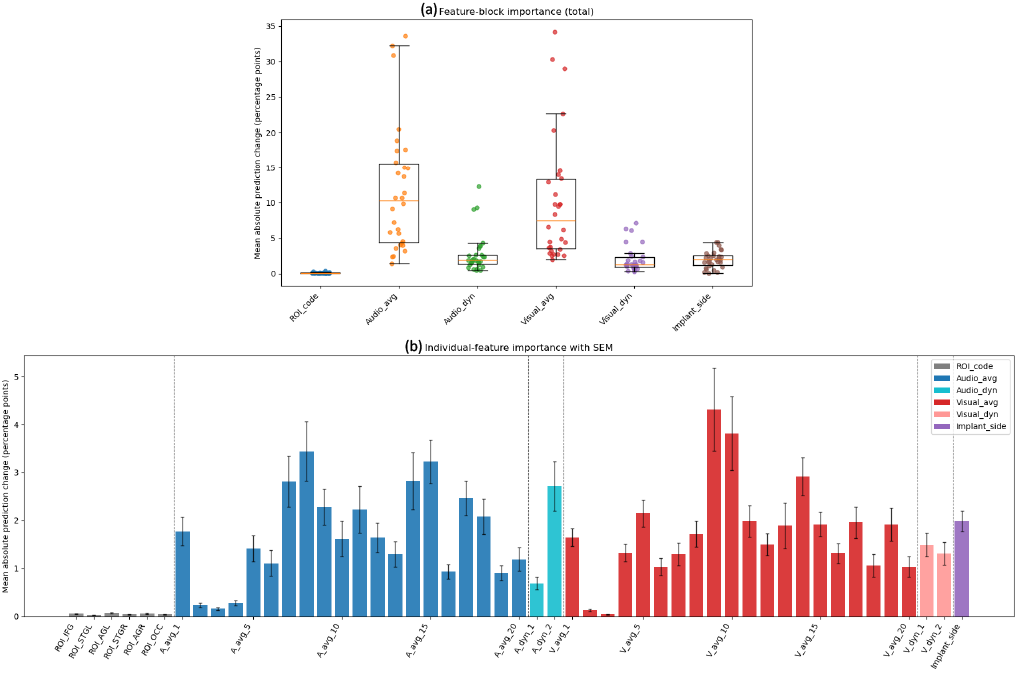
Node-level feature importance analysis using perturbation-based evaluation. (a) Feature-block importance, quantified as the mean absolute change in predicted outcome following perturbation of each feature group. (b) Individual-feature importance with standard error of the mean (SEM), illustrating the contribution of each temporal and dynamic feature.

At the feature-block level (Figure 4a), the model exhibited markedly higher sensitivity to waveform-derived features compared to other inputs. Specifically, both auditory and visual waveform features (average responses) contributed most strongly to prediction performance, followed by their corresponding dynamic features. The implant side also showed a non-negligible contribution, with an importance comparable to that of the dynamic features. In contrast, ROI encoding had minimal influence on the model output. While the higher importance of waveform features is partly attributable to their greater dimensionality, a key observation is that auditory and visual modalities contribute at comparable levels across both average and dynamic representations, with no significant differences observed between corresponding auditory and visual feature groups (waveform: 11.8% vs. 11.6%, *p* = 0.919; dynamic: 2.8% vs. 2.0%, *p* = 0.225). This suggests that the model integrates complementary information from both auditory and visual processing streams, rather than relying on a single dominant modality. At the same time, the contribution of implant side indicates that clinically relevant non-imaging information provides additional predictive value when combined with brain-derived features.

At the individual-feature level (Figure 4b), the temporal distribution of feature importance for average waveforms closely follows the expected hemodynamic response profile. Peak importance is observed at delayed time points corresponding to maximal vascular responses [32], whereas early time points contribute minimally. This indicates that the model preferentially utilizes physiologically meaningful components of the fNIRS signal, rather than time points dominated by baseline activity or noise. Consistent with the preprocessing pipeline, time samples near stimulus onset (second and third samples) exhibited near-zero importance, likely reflecting reduced variability following baseline correction. This temporal selectivity further supports that the model captures task-relevant neural responses rather than spurious signal fluctuations.

Furthermore, ROI identity—encoded as one-hot vectors—showed negligible contribution to prediction performance. This suggests that explicit anatomical labeling provides limited additional information beyond what is already captured by the functional characteristics of the signals and their interactions within the graph structure.

### E. Aggregation-Level Node and Edge Importance

To further examine the role of higher-level graph components, we evaluated the importance of aggregation nodes and inter-aggregation edges using the same perturbation-based framework (Figure 5).

**Fig. 5.**
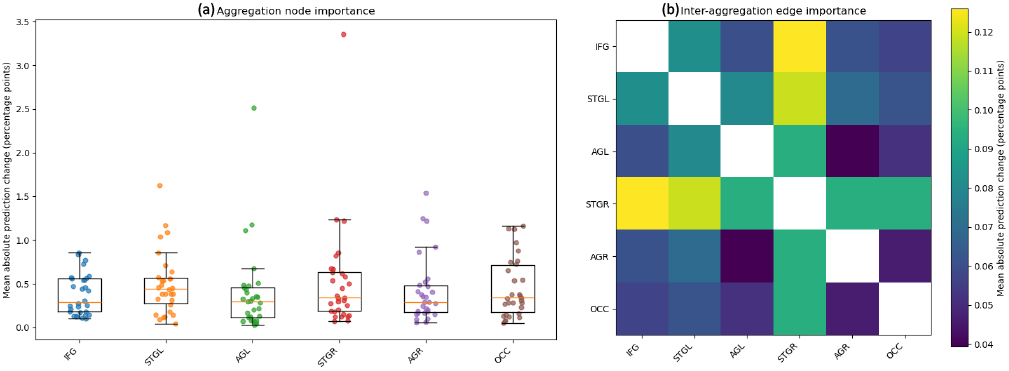
Aggregation-level node and edge importance analysis. (a) Importance of ROI-level aggregation nodes and (b) Heatmap of inter-aggregation edge importance, representing the mean absolute change in prediction following perturbation of the components.

At the aggregation node level (Figure 5a), no clear or consistent differences were observed across ROIs. All regions exhibited comparable importance values, with no significant effect of ROI on aggregation-node importance (Friedman test: *χ*^2^(5) = 6.34, *p* = 0.274), suggesting that no single aggregation node dominates the prediction. This indicates that outcome prediction relies on distributed contributions across regions rather than a specific ROI acting in isolation.

In contrast, the analysis of inter-aggregation edges (Figure 5b) revealed more structured patterns. Connections involving the right superior temporal gyrus (STGR) aggregation node showed consistently higher importance compared to other inter-regional links. In particular, edges connecting STGR with frontal and other temporal regions contributed most strongly to the model’s predictions.

## IV. Discussion

The present study introduces a graph-based multivariable framework for predicting cochlear implant outcomes from early post-implantation fNIRS recordings. By integrating multiple aspects of brain organization—including regional activation (segregation), inter-regional interactions (integration), modular structure, and temporal dynamics—the proposed model provides a unified representation of brain function that significantly outperforms previously reported single-biomarker approaches [28]–[30]. These findings support the central premise that clinically relevant brain behavior is best understood as a distributed and multivariate phenomenon rather than through isolated neural indicators [1], [3], [5].

A key contribution of this work lies in bridging two traditionally distinct paradigms in brain imaging analysis. Classical approaches can broadly be divided into (i) activation-based methods, such as general linear modeling (GLM) or waveform averaging, which focus on regional responses to stimuli [33], and (ii) network-based approaches grounded in graph theory, which characterize interactions between brain regions [4]. These two perspectives are often studied separately, despite representing complementary aspects of brain function. The proposed GNN-based framework provides a natural platform to integrate both. In this formulation, brain activation patterns are encoded as node features, capturing localized responses, while the graph structure represents interactions between regions. This unified representation enables simultaneous modeling of regional activity and network-level organization within a single predictive framework.

More fundamentally, GNNs offer a principled way to model brain data that departs from traditional Euclidean-based machine learning approaches. Unlike convolutional or perceptron neural networks, which operate on regular grid structures, GNNs are designed to process data defined on irregular domains such as graphs [34]. In this context, brain networks are naturally represented as nodes (regions or channels) connected by edges (interactions). Through iterative message-passing operations, each node updates its representation by aggregating information from its neighbors. After multiple layers of such updates, node embeddings reflect not only local features but also the structural and functional context of their surrounding network. This allows the model to capture complex, non-local dependencies and higher-order interactions that are difficult to represent using conventional approaches [25].

A key implication of this work is the limitation of hypothesis-driven, univariate approaches in capturing the complexity of brain function. While such methods have provided valuable insights into specific mechanisms, they inherently rely on prior assumptions about which features are most relevant. In many translational settings, however, the full set of factors influencing brain behavior—whether developmental, adaptive, or pathological—remains unknown. As a result, restricting analysis to predefined biomarkers risks overlooking important contributing factors and interactions. In contrast, hypothesis-free multivariable models allow the data to inform the representation, enabling the identification of distributed patterns that may not be captured by conventional approaches. This is particularly important in clinical prediction tasks, where the goal is to achieve robust and generalizable estimation of individual outcomes.

Beyond predictive performance, multivariable frameworks offer an additional advantage: they can serve as tools for discovery. When combined with exploratory analyses, such as the perturbation-based approach employed in this study, these models can provide insights into the relative importance of different components of brain function. For example, our findings highlight the role of interactions involving the STGR as influential contributors to prediction. While this observation should be interpreted cautiously, it is broadly consistent with evidence suggesting a distinct role of STGR in auditory processing and multisensory integration. In the context of cochlear implantation, this may reflect the involvement of STGR in adapting to degraded auditory input and integrating information across modalities [35], [36]. More broadly, such observations illustrate how multivariable models can generate hypotheses and guide future investigations, complementing traditional hypothesis-driven research.

Another important aspect of the proposed framework is its explicit incorporation of modular organization. The brain is known to exhibit hierarchical modularity, where functional systems are organized into nested structures across multiple spatial scales [37], [38]. In this study, we modeled a two-level hierarchy consisting of channel-level nodes and ROI-level aggregation nodes, enabling representation of both intraregional and inter-regional interactions. This design provides a balance between model expressiveness and interpretability, allowing the learned representations to remain closely aligned with known anatomical and functional organization. Notably, the relatively compact graph structure used in this work may also contribute to improved robustness and interpretability compared to large-scale fully-connected graphs, where noise and redundancy can obscure meaningful patterns.

An additional advantage of the proposed framework becomes particularly evident when considering higher-resolution imaging modalities such as fMRI. In such settings, dimensionality is often reduced by summarizing voxel-level signals using averaging, principal component analysis, or independent component analysis [39], [40]. While effective for reducing complexity, these approaches may discard fine-grained spatial information and impose assumptions about the underlying structure of the data. In contrast, the graph-based formulation adopted here provides a flexible framework for representing brain organization across multiple spatial scales while preserving local information within a structured hierarchy. This enables the integration of fine-grained and large-scale neural processes without relying on aggressive dimensionality reduction, offering a balance between representational richness and interpretability in high-dimensional neuroimaging data.

### A. Future Directions

Several avenues exist for extending and improving the proposed framework. First, the modeling of temporal dynamics can be further refined. While the current approach employs compact features to capture trial-to-trial variability, we also explored more explicit representations of temporal structure, including higher-dimensional trial-level modeling. In particular, we investigated hybrid architectures that combine convolutional neural networks with graph-based models, motivated by recent work on waveform-level decoding of fNIRS responses (manuscript under revision following peer review by IEEE Trans. Neural Syst. Rehabil. Eng.). However, these approaches did not consistently improve predictive performance, likely due to increased model complexity relative to the available sample size and variability across trials. Consequently, a more parsimonious representation was adopted by engineering two simple yet physiologically meaningful features to summarize key aspects of trial-to-trial dynamics while maintaining model stability. Future studies with larger datasets or alternative architectures—such as sequence-based models or attention mechanisms operating across trials—may enable more comprehensive and robust modeling of temporal dynamics. In addition, given the limited contribution of explicit ROI encoding observed in this study, future work may explore alternative feature representations that reduce redundancy while preserving spatial and functional information.

Second, the framework can be extended to incorporate multi-level hierarchical modularity, particularly for high-resolution imaging modalities such as fMRI. As illustrated by hierarchical clustering approaches and the proposed two-level modular graph design in Figure 6, brain networks can be represented across multiple spatial scales, ranging from fine-grained local modules to higher-order functional systems. In this representation, lower-level modules are aggregated into higher-level structures via intermediate aggregation nodes, enabling a hierarchical organization of the network. For higher-resolution data, this approach can be naturally extended by introducing additional levels of aggregation, resulting in deeper hierarchical graphs. Correspondingly, the GNN architecture can be expanded with additional layers to propagate information across these levels, allowing the model to capture both local dynamics and large-scale inter-module interactions within a unified framework.

**Fig. 6.**
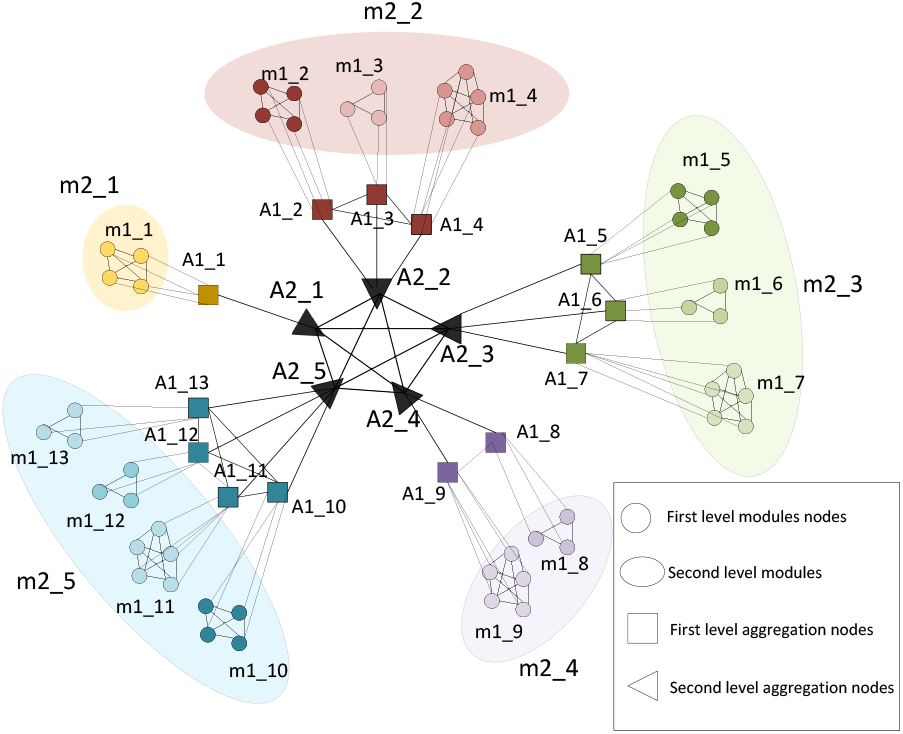
Two-level hierarchical graph representation of brain networks. Channel-level nodes (small circles, labeled ***m*1_*i***) are grouped into first-level modules (labeled ***m*2_*i***), representing localized functional units. Each first-level module is summarized by an aggregation node (***A*1_*i***, squares), which captures the collective information of its constituent nodes. These first-level modules are further organized into second-level modules (labeled ***m*2_*i***), reflecting higher-order functional groupings. Second-level aggregation nodes (***A*2_*i***, triangles) integrate information across first-level modules and are interconnected to model large-scale network interactions.

## V. Conclusion

In this study, we proposed a graph-based multivariable framework for predicting cochlear implant outcomes from early post-implantation fNIRS recordings. By integrating multiple aspects of brain organization—including regional activation, inter-regional interactions, modular structure, and temporal dynamics—within a unified GNN architecture, the model provides a comprehensive representation of brain function. The results demonstrate that this multivariable approach achieves substantially improved predictive performance compared to traditional single-biomarker methods, highlighting the limitations of univariate analyses in capturing the distributed nature of neural processes. Beyond predictive accuracy, the framework offers valuable insights into the mechanisms underlying outcome variability. Perturbation-based analyses indicate that prediction is driven by distributed functional patterns across modalities and regions, rather than isolated features, reinforcing the importance of modeling brain function as a network-level phenomenon. The identification of specific inter-regional interactions further illustrates the potential of multivariable models to generate new hypotheses about brain organization and adaptation. More broadly, this work demonstrates the utility of graph neural networks as a flexible platform for integrating activation-based and connectivity-based representations within a single model. By explicitly incorporating hierarchical modular structure and multiscale interactions, the proposed approach provides a principled way to model the complexity of brain systems. These characteristics make the framework well suited for extension to other neuroimaging modalities and clinical applications.

## Data Availability

All data produced in the present study are available upon reasonable request to the authors

## Notes

This study was supported by the National Health and Medical Research Council of Australia (2016 Conjoint Grant), a Junior Fellowship from the Passe and Williams Foundation (GNT1163894 to T.P.), and the University of Melbourne Research Scholarship (625441 to J.E.). The Bionics Institute acknowledges the support it receives from the Victorian Government through its Operational Infrastructure Support Program.

### Competing Interest Statement

The authors have declared no competing interest.

### Author Declarations

Ethical approval for the study was obtained from the Royal Victorian Eye and Ear Hospital (approval no. 19.1418H), and all participants provided written informed consent.

